# Plasma neutralization properties of the SARS-CoV-2 Omicron variant

**DOI:** 10.1101/2021.12.12.21267646

**Authors:** Fabian Schmidt, Frauke Muecksch, Yiska Weisblum, Justin Da Silva, Eva Bednarski, Alice Cho, Zijun Wang, Christian Gaebler, Marina Caskey, Michel C. Nussenzweig, Theodora Hatziioannou, Paul D. Bieniasz

## Abstract

**BACKGROUND:** The Omicron SARS-CoV-2 variant has spread internationally and is responsible for rapidly increasing case numbers. The emergence of divergent variants in the context of a heterogeneous and evolving neutralizing antibody response in host populations might compromise protection afforded by vaccines or prior infection.

**METHODS:** We measured neutralizing antibody titers in 169 longitudinally collected plasma samples using pseudotypes bearing the Wuhan-hu-1 or the Omicron variant or a laboratory-designed neutralization-resistant SARS-CoV-2 spike (PMS20). Plasmas were obtained from convalescents who did or did not subsequently receive an mRNA vaccine, or naive individuals who received 3-doses of mRNA or 1-dose Ad26 vaccines. Samples were collected approximately 1, 5-6 and 12 months after initial vaccination or infection.

**RESULTS:** Like PMS20, the Omicron spike protein was substantially resistant to neutralization compared to Wuhan-hu-1. In convalescent plasma the median deficit in neutralizing activity against PMS20 or Omicron was 30- to 60-fold. Plasmas from recipients of 2 mRNA vaccine doses were 30- to 180- fold less potent against PMS20 and Omicron than Wuhan-hu-1. Notably, previously infected or two-mRNA dose vaccinated individuals who received additional mRNA vaccine dose(s) had 38 to 154-fold and 35 to 214-fold increases in neutralizing activity against Omicron and PMS20 respectively.

**CONCLUSIONS:** Omicron exhibits similar distribution of sequence changes and neutralization resistance as does a laboratory-designed neutralization-resistant spike protein, suggesting natural evolutionary pressure to evade the human antibody response. Currently available mRNA vaccine boosters, that may promote antibody affinity maturation, significantly ameliorate SARS-CoV-2 neutralizing antibody titers.

## Introduction

The recent emergence of the B.1.1.159 (Omicron) variant of SARS-CoV-2 ^1-3^ has engendered widespread concern. Omicron has spread internationally and is responsible for rapidly increasing case numbers, particularly in South Africa^3^. Although the pathogenic potential of the new variant remains unclear^3^, a striking feature is the large number of amino acid substitutions, insertions and deletions in the spike protein (32 of the total of ∼50 nonsynonymous changes in the viral genome)^1^ suggesting adaptation to substantial selective pressure. Some of the substitutions, for example those proximal to the furin cleavage site, are thought to be fitness enhancing and may facilitate virus spread^4^, but the majority of the changes are expected to reduce neutralizing antibody recognition.

The number of neutralizing epitopes targeted by polyclonal antibodies in SARS-CoV-2 convalescent or vaccinated individuals is an important determinant of the genetic barrier to viral escape^5^. Whereas single monoclonal antibodies are prone to escape mutations, combinations targeting non-overlapping epitopes are more resistant to such changes^6,7^. There are numerous antibody targets in the SARS-CoV-2 spike protein, but polyclonal neutralizing responses are dominated by antibodies to the receptor binding domain (RBD) and the N-terminal domain (NTD) of spike^5,8-10^. Indeed, aggregation of ∼20 RBD and NTD mutations in a polymutant spike protein (PMS20) was required for evasion of polyclonal antibodies elicited in the majority of individuals who had been infected, or who had received two doses of an mRNA vaccine^5,15^. Notably, several of the changes in the PMS20 spike are the same or similar to the changes in the emergent Omicron variant spike^11,12^ (Figure 1A, B), leading to the prediction that Omicron would exhibit substantial antigenic escape.

**Figure 1.**
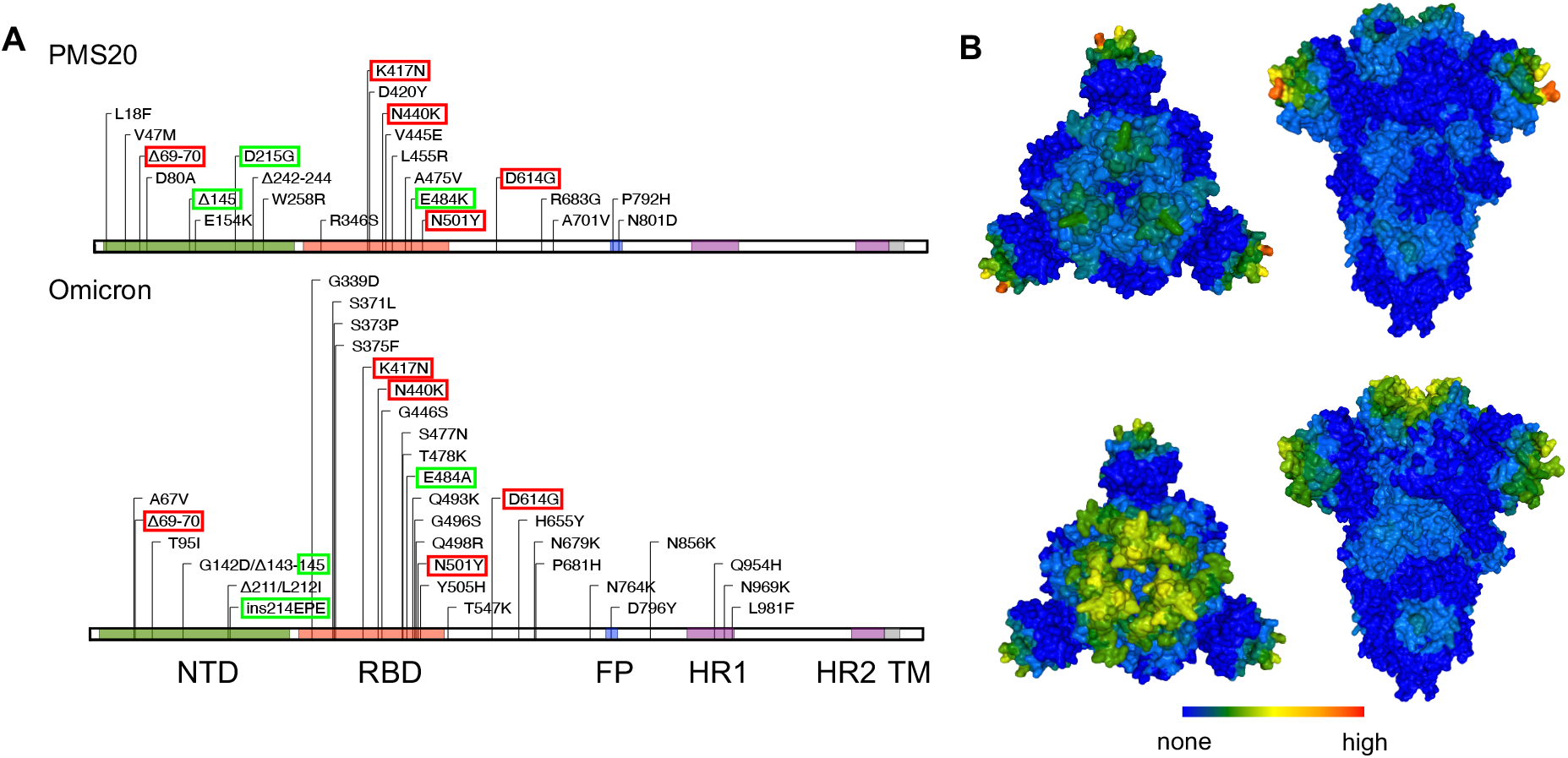
Distribution of mutations in the naturally emergent Omicron variant, and the neutralization resistant designed PMS20 spike proteins. (A) Schematic diagram of PMS20 and Omicron spike proteins with amino acid changes compared to Wuhan-1 indicated. Red outlines highlight identical changes in PMS20 and Omicron, green outlines indicate nonidentical changes affecting same residues in PMS20 and Omicron. Position of spike features is marked: NTD = N-terminal domain RBD = Receptor Binding Domain, FP = Fusion Peptide, HR = heptad repeat, TM = Transmembrane. (B) Representation of the spatial distribution of mutations on the surface of PMS20 and Omicron spike proteins. Frequency of amino acid changes relative to Wuhan-hu-1 in PMS20 or Omicron projected onto the SARS-CoV-2 spike structure (PDB 6VXX). The average frequency of substitutions in a 15 angstrom radius is represented using a color spectrum (scale 0-40) (https://biostructmap.burnet.edu.au).

Affinity maturation of individual SARS-CoV-2 neutralizing antibodies can dramatically alter their properties in ways that are pertinent for the emergence and control of variants^13-16^. The number of antibody variable region mutations and the binding affinity of antibodies increases over months and can vary depending on the nature of SARS-CoV-2 antigen exposure^13,15-18^. Indeed, affinity maturation can markedly expand SARS-CoV-2 neutralizing antibody breadth, enabling neutralization of SARS-CoV-2 variants that escape neutralization by corresponding ancestral antibodies and imposing a requirement for multiple amino acid substitutions for escape^7,13-15,18,19^. Thus, Omicron has emerged in the context of a globally heterogeneous and evolving neutralizing antibody response in host populations that might provide varying degrees of protection, depending on infection and vaccination history. Here, we determined the ability of individuals with varying exposure to SARS-CoV-2 infection and vaccination, to neutralize SARS-CoV-2 pseudotypes with spike proteins corresponding to the parental virus used in vaccine immunogens, PMS20 or the emergent Omicron variant.

## Methods

### PLASMA SAMPLES

The 169 plasma samples were from the following three longitudinal cohorts (Table 1): (i) Convalescent individuals who did or did not receive 2 doses of the Pfizer/BNT or Moderna mRNA vaccine between 6 months and 12 months after infection^9,13,15^; (ii) Uninfected individuals who received 3 doses of the Pfizer/BNT mRNA vaccine^20^; (iii) Uninfected individuals who received the J&J Ad26 vaccine. Plasma samples were collected approximately 1, 5-6 and 12 months after initial vaccination or infection and selected for this study at random with respect to binding, neutralization titer, or donor demographic characteristics. The study visits and blood draws were reviewed and approved by the Institutional Review Board of the Rockefeller University (IRB no. DRO-1006, ‘Peripheral Blood of Coronavirus Survivors to Identify Virus-Neutralizing Antibodies’).

**Table 1.** Plasma samples tested for neutralizing activity.

| Plasma panel | *N* | Description |  | Time after infection or vaccination |
| --- | --- | --- | --- | --- |
| Panel 1<sup>1</sup> | 20 | Convalescent |  | ~1m post infection |
| Panel 2<sup>1</sup> | 20 | Convalescent |  | ~6m post infection |
| Panel 3 | 20 | Convalescent |  | ~1y post infection |
| Panel 4<sup>2</sup> | 18 | Vaccinated | 2 doses of Pfizer mRNA | ~1m post 2<sup>nd</sup> dose |
| Panel 5<sup>2</sup> | 18 | Vaccinated | 2 doses of Pfizer mRNA | ~5m post 2<sup>nd</sup> dose |
| Panel 6<sup>2</sup> | 18 | Vaccinated | 3 doses of Pfizer mRNA | ~1m post 3<sup>rd</sup> dose |
| Panel 7<sup>3</sup> | 19 | Vaccinated | 1 dose J&J Ad26 | ~1m post single dose |
| Panel 8<sup>3</sup> | 19 | Vaccinated | 1 dose J&J Ad26 | ~5m post single dose |
| Panel 9<sup>1</sup> | 17 | Convalescent<br>+ Vaccinated | 1 or 2 doses of Pfizer or Moderna mRNA, following prior SARS-CoV-2 infection | ~12m post infection, various times post vaccination |

<sup>1</sup>Plasma panels 1, 2, and 9 from matched convalescent individuals
<sup>2</sup>Plasma panels 4, 5, and 6 from matched mRNA vaccinated individuals
<sup>3</sup>Plasma panels 7 and 8 from matched J&J Ad26 vaccinated individuals

### PSEUDOTYPE NEUTRALIZATION ASSAY

The Omicron spike coding sequence was derived from sequence ID EPI_ISL_6640919. It was codon-optimized and synthesized as a C-terminally truncated Δ19 form in nine fragments (IDT). We also introduced a furin cleavage site mutation (R683G) that does not change the neutralization properties of the SASR-CoV-2 spike protein but enables higher titer pseudotyped viral stocks to be generated from transfected cells^5^. These synthetic DNA fragments, ranging in size from 444-599bp and a NheI/XbaI-linearized pCR3.1 plasmid were Gibson assembled via 40bps overlapping sequences. Individual plasmid clones were completely sequenced (Illumina MiSeq) and a single correct clone was used in these studies. The Wuhan-hu-1 and PMS20 spike proteins were previously described^5,21^.

Neutralizing titers were measured using a SARS-CoV-2 pseudotyped HIV-1-based assay that recapitulates neutralizing titers obtained with authentic SARS-CoV-2^21^. Plasmas were serially diluted (five-fold dilution interval) and then incubated with a SARS-CoV-2 spike (Wuhan-hu-1, PMS20 or Omicron) pseudotyped HIV-1 based nanoluc luciferase reporter virus for 1 hour at 37 °C. The pseudotyped virus and antibody mixture was transferred to 96 well plates containing HT1080/ACE2.cl14 cells. After 48 hours, the cells were washed with PBS and lysed with Luciferase Cell Culture Lysis reagent (Promega). Then Nanoluc Luciferase activity in cell lysates was measured using the Nano-Glo Luciferase Assay System and a Glomax Navigator luminometer (Promega). The relative luminescence units were normalized to those measured in cells infected with the corresponding pseudotyped virus in the absence of plasma. The half-maximal plasma neutralizing titer (NT_50_) was determined using four-parameter nonlinear regression (least squares regression method without weighting) (GraphPad Prism). The NT50 for each plasma was measured twice in two independent experiments, carried out by two different groups of researchers, with two technical replicates each.

## Results

We measured plasma neutralizing antibody titers in 169 plasma samples from 47 individuals who have experienced diverse exposures to SARS-CoV-2 antigens (Table 1). Specifically, we measured titers of convalescent individuals who had been infected early in the pandemic, and whose plasma was donated an average of 1.3 and 6.2 months after infection (Supplementary Table S1). These individuals subsequently received one or two doses of an mRNA vaccine (Pfizer/BNT or Moderna) before collection of additional plasma samples approximately 12 months after initial infection. For comparison, we measured neutralizing titers in a group of convalescent individuals from the same initial cohort who were not vaccinated (Supplementary Table S1). We also measured neutralizing titers in uninfected individuals that received 2 doses of an mRNA vaccine (Pfizer/BNT or Moderna) and donated plasma at 1.2 and 5 months after the second dose. These same individuals received a third Pfizer/BNT dose booster >6 months after the second dose and donated another plasma sample 1 month after the third dose (Supplementary Table S2). Finally, we examined a cohort that received an Ad26 adenovirus vaccine (J&J) that has been widely deployed in the USA, at ∼1 and 5 months after vaccination (Supplementary Table S2).

### NEUTRALIZATION OF PMS20 AND OMICRON PSEUDOTYPES BY CONVALESCENT PLASMA

We found that, like PMS20, the Omicron variant pseudotyped virus was substantially resistant to neutralization by plasma compared to Wuhan-hu-1. In convalescent individuals whose plasmas were collected at ∼1.2 months after infection, the median and range of the NT_50_ values was 2616 (681-19450), 49 (<25-429) and 68 (29-667) for the Wuhan-hu-1, PMS20 and Omicron spike proteins respectively (Figure 2A). These values represent a mean (±SD) fold reduction in NT_50_ of 60±47 -fold for PMS20 and 58±51-fold for Omicron compared to Wuhan-hu-1. After 6 months of convalescence, the median and range of NT_50_ values from the same individuals was 1678 (321 - 5189), 28 (<25 - 248), and 42 (<25 - 428) for the Wuhan-hu-1, PMS20 and Omicron spike proteins respectively, corresponding to mean (±SD) reductions in NT_50_ of 37±27-fold for PMS20 and 32±23-fold for Omicron (Figure 2B). Similarly, plasmas collected from different individuals in the same cohort, 1 year after infection had NT_50_ values (median and range) of 2037 (127 - 25835), 76 (<25 - 907) and 136 (<25 - 667) for Wuhan-hu-1, PMS20 and Omicron pseudotypes, i.e. mean (±SD) reductions in NT_50_ of 34±24-fold for PMS20 and 43±23-fold for Omicron (Figure 2C). Overall, individuals who were infected but not vaccinated show substantially reduced plasma neutralizing activity including many with sub detectable titers against Omicron pseudotypes.

**Figure 2.**
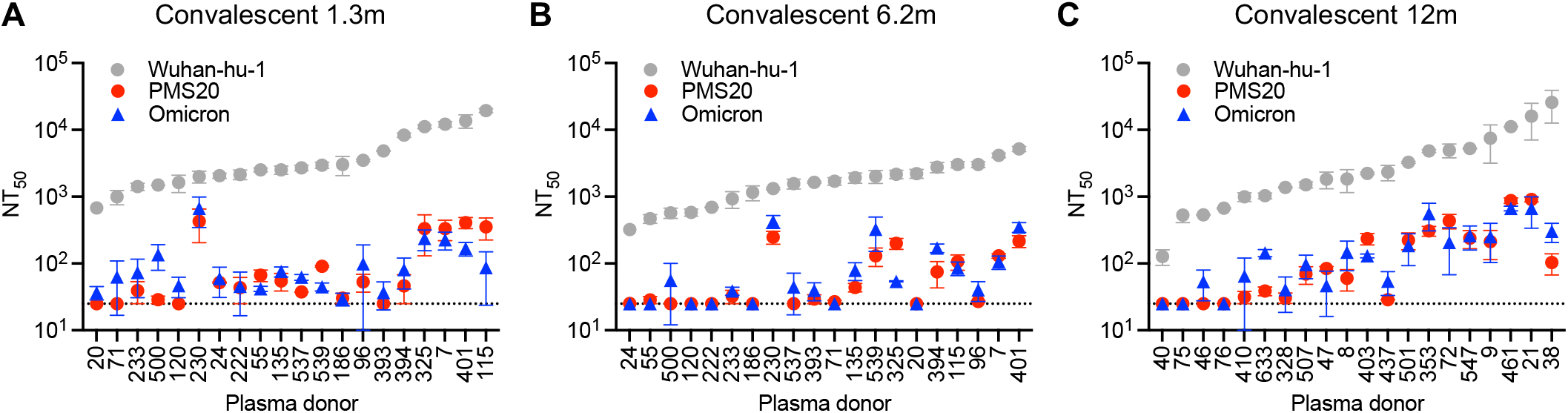
Plasma neutralizing titers against Wuhan-hu-1, PMS20 and Omicron SARS-CoV-2 variants in convalescent, unvaccinated individuals. (A-C) NT_50_ values of plasmas collected at 1 month (A), 6 months (B) and 12 months (C) after SARS-CoV-2 infection against Wuhan-hu-1, PMS20 and Omicron spike pseudotyped reporter viruses. Each NT_50_ was determined in two independent experiments (each with two technical replicates). The median and range of the two independent determinations is plotted. Dashed line indicates the lowest plasma dilution tested (1:25).

### NEUTRALIZATION OF PMS20 AND OMICRON PSEUDOTYPES BY VACCINE RECIPIENT PLASMA

Plasmas from vaccinated individuals also exhibited substantially impaired ability to neutralize the Omicron variant. In plasmas from individuals who received mRNA vaccines ∼1.3m prior to sampling, the median and range of the NT_50_ values were 7627 (2299 - 50640), for Wuhan-hu-1, but only 60 (<25 - 201) for PMS20 and 92 (25 - 327) for Omicron (Figure 3A). These values correspond to mean (±SD) loss of potency of 187±24 fold for PMS20 and 127±66 fold for Omicron. At 5 months after vaccination, neutralizing titer (median and range) in plasmas from the same individuals had waned to 2435 (1117 – 6228) for Wuhan-hu-1 and were as low as 43 (<25 – 108) for PMS20 and 126 (27 – 321) for Omicron, corresponding to mean (±SD) reductions in NT_50_ of 58±23-fold for PMS20 and 27±17-fold for Omicron (Figure 3B).

**Figure 3.**
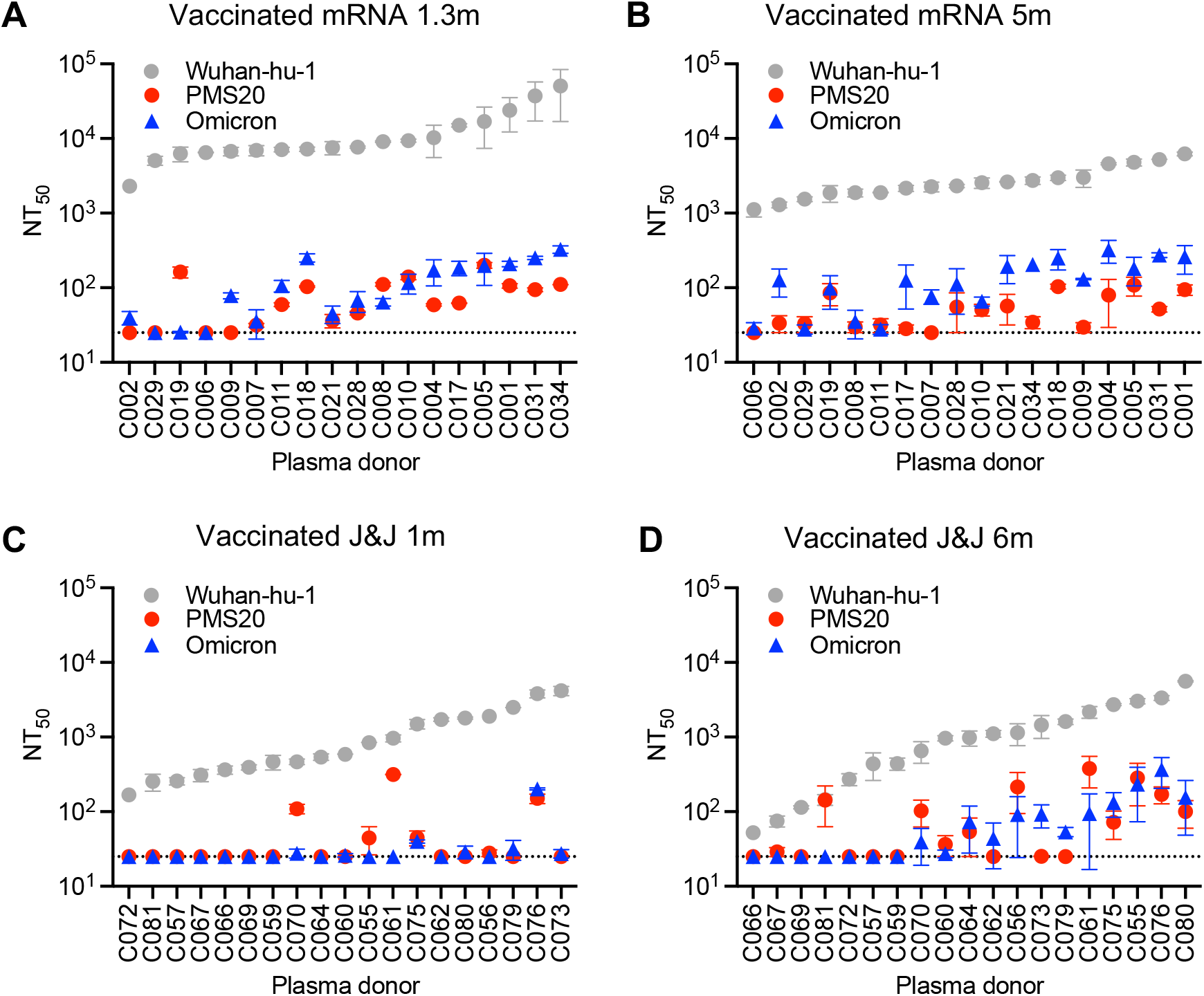
Plasma neutralizing titers against Wuhan-hu-1, PMS20 and Omicron SARS-CoV-2 variants in vaccine recipients. (A, B) NT_50_ values of plasmas from recipients of standard 2 dose mRNA (Pfizer/BNT or Moderna) approximately one (A) and five (B) months after vaccination. (C,D) NT_50_ values of plasmas from recipients of single dose Ad26 (J&J) SARS-CoV-2 vaccines approximately one (C) and five (D) months after vaccination. For each panel, NT_50_ were determined in two independent experiments (each with two technical replicates). The median and range of the two independent determinations is plotted. Dashed line indicates the lowest plasma dilution tested (1:25)

Plasmas from J&J vaccine recipients were particularly poor at neutralizing the variants. Plasma from J&J vaccine recipients collected 1 month after vaccination had median (and range) NT_50_ values of 588 (167 - 4177), <25 (<25-314) and <25 (<25-201) for the Wuhan-hu-1, PMS20 and Omicron spike proteins respectively (Figure 3C). NT_50_ values for J&J recipients were quite stable or even increased in some cases over time, such that at five months after J&J vaccination NT_50_ values were 982 (52 – 5597), 36 (25 – 378) and 43 (<25 – 368) for the Wuhan-hu-1, PMS20 and Omicron pseudotypes respectively (Figure 3D). Nevertheless, neutralizing titers in the J&J vaccinees were sufficiently low, such that quantitative reductions in potency against the PMS20 and Omicron pseudotypes could not be meaningfully calculated because many plasmas did not contain detectable neutralizing activity against these pseudotypes (Figure 3B).

### EFFECTS OF mRNA VACCINE BOOSTERS ON NEUTRALIZATION BY CONVALESCENT OR PREVIOUSLY mRNA VACCINATED INDIVIDUALS

Vaccination of previously infected individuals has previously been shown to substantially elevate neutralizing titers and breadth^5,15,22,23^. Notably, vaccination of convalescent individuals^5,15^ or boosting of an initial vaccine response by administration of a third mRNA (Pfizer/BNT) vaccine dose >6 months after the initial series led to the acquisition of substantial neutralizing activity against PMS20 and against Omicron (Figure 4A, B). Specifically, in individuals that were previously infected by SARS-CoV-2 and later vaccinated, the median and range of the NT_50_ values was 388872 (98522 - 1304453), 14982 (1699 - 448699) and 8106 (1503 - 56537) for the Wuhan-hu-1, PMS20 and Omicron spike protein pseudotypes respectively (Figure 4A). Analysis of NT_50_ trajectories for individual vaccine recipients showed that these values represent a median increase in NT_50_ of 238-fold, 214-fold and 154-fold for Wuhan-hu-1, PMS20 and Omicron pseudotypes compared to the pre-vaccination, convalescent titers in the same individuals (Figure 4C).

**Figure 4.**
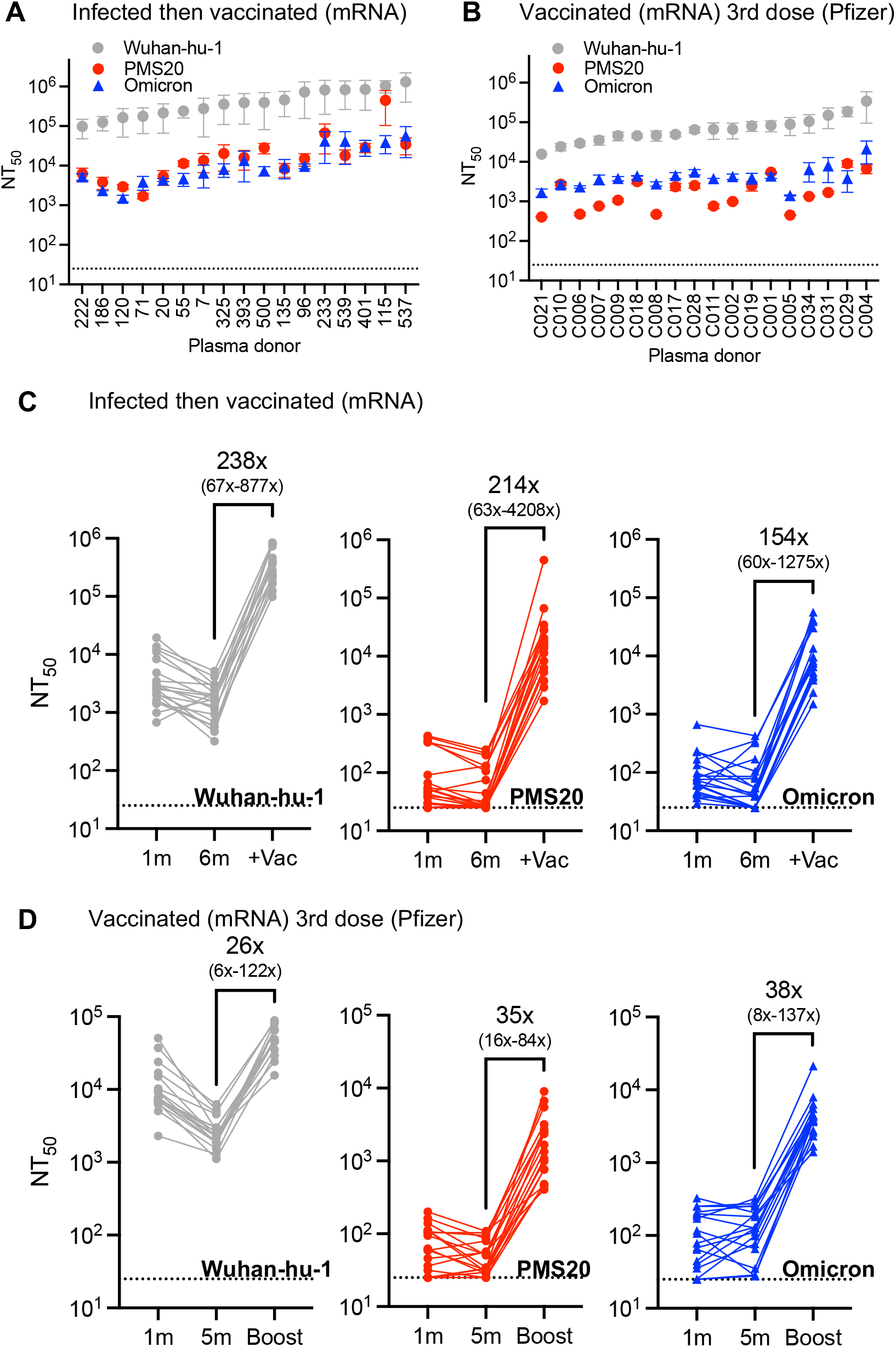
Effect of vaccinating previously infected individuals or boosting previously vaccinated individuals on Wuhan-hu-1, PMS20 and Omicron plasma neutralizing titers. (A) NT_50_ values of plasmas from recipients of mRNA (Pfizer/BNT or Moderna) vaccines subsequent to infection with SARS-CoV-2. (B) NT_50_ values of plasmas from recipients of three doses of mRNA (Pfizer/BNT or Moderna) vaccines with the third dose (Pfizer/BNT) administered >6m after the second. For (A) and (B) each NT_50_ was determined in two independent experiments (each with two technical replicates). The median and range of the two independent determinations is plotted. Dashed line indicates the lowest plasma dilution tested (1:25). (C) Trajectories of NT_50_ values against Wuhan-hu-1, PMS20, and Omicron pseudotypes, as indicated, for convalescent individuals measured 1 month and 6 months after infection and then following subsequent vaccination (+Vac). (D) Trajectories of NT_50_ values against Wuhan-hu-1, PMS20, and Omicron pseudotypes, as indicated, for mRNA vaccinated individuals measured 1 month and 5 months after the second dose and 1 month after the third dose (Boost).

For individuals that were vaccinated with 2 doses of a Pfizer/BNT or Moderna mRNA vaccines ∼6 months previously and then boosted with a third mRNA (Pfizer/BNT) dose ∼1month prior to sampling, the median and range of the NT_50_ values was 65617 (15641 - 341247), 1505 (404 - 9039) and 3871 (1411 - 21300) for the Wuhan-hu-1, PMS20 and Omicron pseudotypes, representing a comparative deficit in potency of 55±45 fold for PMS20 and 18±14 fold for Omicron (Figure 4B). Nevertheless, compared to NT_50_ values measured at 5 months after the second mRNA vaccine dose (prior to the booster) in the same individuals, these boosted titers represent a median (+/-SD) NT_50_ increase of 26-fold for Wuhan-hu-1, 35-fold for PMS20 and 38-fold for Omicron (Figure 4D). We conclude that boosting immunity with mRNA vaccines dramatically increases neutralizing titer and breadth for individuals who had previously been infected with Wuhan-hu-1-like SARS-CoV-2 or vaccinated with Wuhan-hu-1-based mRNA vaccines. In particular, neutralizing titers against Omicron were substantial (1411-56537) in all infected then vaccinated or three dose mRNA vaccine recipients tested, but low or undetectable in many convalescent unvaccinated or two dose mRNA vaccine recipient individuals (Figure 4C, D).

## Discussion

Compared with previous naturally occurring SARS-CoV-2 variants, Omicron exhibits an unprecedented degree of neutralizing antibody escape. Indeed, the degree of neutralization resistance exhibited by Omicron is similar to that exhibited by PMS20, a designed neutralization resistant spike in which 20 naturally occurring and laboratory selected mutations were aggregated^5^. The similarity in neutralization properties and distribution of changes on the spike protein surface between PMS20 and Omicron argues that a major selective pressure leading to the emergence of Omicron was imposed by neutralizing antibodies. Whether this selective pressure occurred in one or more immunocompromised individuals with persistent infection, or populations that have experienced high prevalence infection by waves of prior variants remains unclear.

Of particular concern, neutralizing antibody titers against Omicron were low, even below the limit of detection in a significant fraction of convalescent individuals, Ad26 vaccine recipients or 2 dose mRNA vaccine recipients, particularly following the waning that ensues following infection or vaccination. Nevertheless, individuals that had been previously infected with SARS-CoV-2 and subsequently received mRNA vaccines, or those that have received three doses of mRNA vaccines had substantial neutralizing antibody titers against Omicron ∼1month after boosting. The ability of plasmas from these individuals to neutralize Omicron likely represents the combined effect of increased antibody levels following multiple exposures to antigen, as well as the effects of affinity maturation that can dramatically improve the neutralizing breadth of individual SARS-CoV-2 antibodies as well as polyclonal plasma^5,15^.

These findings suggest that boosting and promoting affinity maturation of the antibodies of those who have previously been infected or vaccinated using existing Wuhan-hu-1 based vaccine immunogens will provide additional protection against Omicron variant infection and disease. The emergence of the Omicron lineage that is phylogenetically distinct from the previously globally dominant Delta SARS-CoV-2 variant also illustrates the need for continuing vigilance and mitigation of virus transmission. The prior emergence, and global spread of the Alpha, Delta, and Omicron variants underscore our inability to predict the emergence of variants that might facilitate targeted vaccine development. The findings described herein and the emergence of Omicron suggest that effort should be devoted to development of vaccination strategies that are broadly based rather than, or in addition to, those narrowly targeted at contemporary emergent SARS-CoV-2 variants.

## Data Availability

All data produced in the present work are contained in the manuscript

## Acknowledgements

This work was supported by NIH grant R37AI64003 and R01AI501111 (P.D.B).; R01AI78788 (TH); P01-AI138398-S1 (M.C.N.) and 2U19AI111825 (M.C.N.). C.G. was supported by the Robert S. Wennett Post-Doctoral Fellowship, in part by the National Center for Advancing Translational Sciences (National Institutes of Health Clinical and Translational Science Award program, grant UL1 TR001866), and by the Shapiro–Silverberg Fund for the Advancement of Translational Research. P.D.B. and M.C.N. are Howard Hughes Medical Institute Investigators.

## Author contributions

P.D.B., T.H., M.C.N., F.S., and F.M. conceived, designed and analyzed the experiments. F.S., Y.W., F.M., J.D.S, and E.B. performed pseudotype neutralization experiments. F.S. constructed expression plasmids. A.C. performed NGS. Y.Z., M.C, C.G. and executed clinical protocols and recruited participants and processed samples. P.D.B., T.H., and M.C.N. wrote the manuscript.

