## Supplemental tables for "Plasma neutralization properties of the SARS-CoV-2 Omicron variant"

This appendix has been provided by the authors to give readers additional information about their work.

**Table S1. Individual convalescent participant characteristics**

| ID | Age (years) | Sex | Temporal dynamics (days): Sx onset to initial visit (1.3m) | Temporal dynamics (days): Sx onset to follow-up visit (1y) | Vaccination status: Vaccine received | Vaccination status: # of doses received prior to 1y | Vaccination status: Days between first dose |
|---|---|---|---|---|---|---|---|
| **8** | 36-40 | M | 57 | 370 | N | N/A | N/A |
| **9** | 31-35 | F | 53 | 366 | N | N/A | N/A |
| **21** | 51-55 | M | 27 | 340 | N | N/A | N/A |
| **38** | 56-60 | F | 38 | 337 | N | N/A | N/A |
| **40** | 41-45 | M | 23 | 345 | N | N/A | N/A |
| **46** | 36-40 | M | 30 | 337 | N | N/A | N/A |
| **47** | 41-45 | F | 33 | 340 | N | N/A | N/A |
| **72** | 41-45 | M | 35 | 352 | N | N/A | N/A |
| **75** | 46-50 | F | 36 | 340 | N | N/A | N/A |
| **76** | 46-50 | F | 34 | 379 | N | N/A | N/A |
| **328** | 51-55 | F | 62 | 365 | N | N/A | N/A |
| **353** | 56-60 | M | 49 | 366 | N | N/A | N/A |
| **403** | 51-55 | M | 39 | 356 | N | N/A | N/A |
| **410** | 31-35 | M | 46 | 349 | N | N/A | N/A |
| **437** | 41-45 | F | 34 | 353 | N | N/A | N/A |
| **461** | 46-50 | M | 39 | 350 | N | N/A | N/A |
| **501** | 31-35 | M | 53 | 367 | N | N/A | N/A |
| **507** | 36-40 | M | 60 | 361 | N | N/A | N/A |
| **547** | 56-60 | M | 36 | 359 | N | N/A | N/A |
| **633** | 36-40 | M | 57 | 358 | N | N/A | N/A |
| **7** | 36-40 | M | 30 | 376 | Pfizer-BNT | 2 | 65 |
| **20** | 26-30 | F | 17 | 345 | Moderna | 2 | 35 |
| **24** | 31-35 | M | 30 | 336 | Pfizer-BNT | N/A | N/A |
| **55** | 36-40 | M | 49 | 349 | Moderna | 2 | 41 |
| **71** | 41-45 | F | 48 | 386 | Pfizer-BNT | 2 | 62 |
| **96** | 46-50 | F | 30 | 359 | Pfizer-BNT | 2 | 54 |
| **115** | 61-65 | F | 41 | 335 | Moderna | 1 | 12 |
| **120** | 56-60 | F | 48 | 375 | Moderna | 2 | 67 |
| **135** | 61-65 | F | 31 | 341 | Moderna | 1 | 27 |
| **186** | 36-40 | F | 33 | 356 | Pfizer-BNT | 2 | 73 |
| **222** | 26-30 | M | 37 | 347 | Pfizer-BNT | 2 | 41 |
| **230** | 46-50 | M | 33 | 372 | Pfizer-BNT | N/A | N/A |
| **233** | 51-55 | M | 41 | 377 | Moderna | 1 | 8 |
| **325** | 51-55 | M | 38 | 353 | Moderna | 2 | 49 |
| **393** | 66-70 | M | 54 | 362 | Moderna | 2 | 57 |
| **394** | 46-50 | F | 67 | 375 | Moderna | N/A | N/A |
| **401** | 61-65 | M | 53 | 371 | Pfizer-BNT | 1 | 18 |
| **500** | 46-50 | M | 53 | 375 | Pfizer-BNT | 1 | 20 |
| **537** | 51-55 | M | 45 | 357 | Pfizer-BNT | 1 | 14 |
| **539** | 71-75 | F | 55 | 362 | Pfizer-BNT | 2 | 50 |

**Table S2. Individual vaccinated participant characteristics**

| ID | Age (years) | Sex | Vaccine platform: 1st dose | Vaccine platform: 2nd dose | Vaccine platform: 3rd dose | Vaccination time line (days): 1st to 2nd dose | 2nd to 3rd dose | 2nd (mRNA)/ 1st (J&J) dose to 1st visit | 2nd (mRNA)/ 1st (J&J) dose to 2nd visit | 3rd (mRNA) dose to Follow-up |
|---|---|---|---|---|---|---|---|---|---|---|
| **C001** | 36-40 | F | Pfizer-BNT | Pfizer-BNT | Pfizer-BNT | 22 | 251 | 35 | 133 | 56 |
| **C002** | 41-45 | M | Pfizer-BNT | Pfizer-BNT | Pfizer-BNT | 21 | 373 | 79 | 177 | 27 |
| **C004** | 36-40 | M | Pfizer-BNT | Pfizer-BNT | Pfizer-BNT | 21 | 389 | 70 | 170 | 27 |
| **C005** | 46-50 | M | Moderna | Moderna | Pfizer-BNT | 28 | 326 | 91 | 175 | 94 |
| **C006** | 61-65 | M | Pfizer-BNT | Pfizer-BNT | Pfizer-BNT | 21 | 246 | 22 | 146 | 22 |
| **C007** | 26-30 | M | Pfizer-BNT | Pfizer-BNT | Pfizer-BNT | 21 | 252 | 37 | 141 | 21 |
| **C008** | 51-55 | F | Pfizer-BNT | Pfizer-BNT | Pfizer-BNT | 21 | 244 | 35 | 141 | 28 |
| **C009** | 31-35 | F | Pfizer-BNT | Pfizer-BNT | Pfizer-BNT | 21 | 244 | 36 | 141 | 22 |
| **C010** | 66-70 | F | Pfizer-BNT | Pfizer-BNT | Pfizer-BNT | 21 | 245 | 34 | 146 | 21 |
| **C011** | 71-75 | F | Moderna | Moderna | Pfizer-BNT | 28 | 274 | 28 | 131 | 21 |
| **C017** | 21-25 | M | Pfizer-BNT | Pfizer-BNT | Pfizer-BNT | 21 | 252 | 21 | 244 | 21 |
| **C018** | 21-25 | F | Pfizer-BNT | Pfizer-BNT | Pfizer-BNT | 21 | 242 | 48 | 172 | 23 |
| **C019** | 31-35 | M | Pfizer-BNT | Pfizer-BNT | Pfizer-BNT | 21 | 245 | 34 | 146 | 21 |
| **C021** | 36-40 | F | Pfizer-BNT | Pfizer-BNT | Pfizer-BNT | 21 | 212 | 28 | 168 | 47 |
| **C028** | 21-25 | M | Pfizer-BNT | Pfizer-BNT | Pfizer-BNT | 21 | 252 | 41 | 168 | 28 |
| **C029** | 56-60 | F | Pfizer-BNT | Pfizer-BNT | Pfizer-BNT | 21 | 244 | 49 | 166 | 22 |
| **C031** | 26-30 | M | Pfizer-BNT | Pfizer-BNT | Pfizer-BNT | 21 | 242 | 37 | 165 | 28 |
| **C034** | 36-40 | F | Pfizer-BNT | Pfizer-BNT | Pfizer-BNT | 24 | 273 | 31 | 245 | 19 |
| **C055** | 36-40 | F | J&J | - | - | - | - | 34 | 175 | - |
| **C056** | 56-60 | F | J&J | - | - | - | - | 52 | 187 | - |
| **C057** | 26-30 | M | J&J | - | - | - | - | 44 | 178 | - |
| **C059** | 21-25 | F | J&J | - | - | - | - | 44 | 181 | - |
| **C060** | 51-55 | M | J&J | - | - | - | - | 35 | 191 | - |
| **C061** | 46-50 | F | J&J | - | - | - | - | 27 | 177 | - |
| **C062** | 31-35 | F | J&J | - | - | - | - | 53 | 200 | - |
| **C064** | 36-40 | F | J&J | - | - | - | - | 46 | 178 | - |
| **C066** | 46-50 | F | J&J | - | - | - | - | 50 | 183 | - |
| **C067** | 46-50 | F | J&J | - | - | - | - | 52 | 193 | - |
| **C069** | 51-55 | F | J&J | - | - | - | - | 37 | 172 | - |
| **C070** | 41-45 | M | J&J | - | - | - | - | 67 | 187 | - |
| **C072** | 41-45 | F | J&J | - | - | - | - | 63 | 150 | - |
| **C073** | 41-45 | F | J&J | - | - | - | - | 56 | 197 | - |
| **C075** | 31-35 | M | J&J | - | - | - | - | 72 | 180 | - |
| **C076** | 31-35 | M | J&J | - | - | - | - | 72 | 179 | - |
| **C079** | 51-55 | M | J&J | - | - | - | - | 46 | 152 | - |
| **C080** | 31-35 | M | J&J | - | - | - | - | 34 | 140 | - |
| **C081** | 41-45 | M | J&J | - | - | - | - | 31 | 136 | - |
